# Knowledge of Pediatric and Adolescent Cancer among Primary Caregivers: An Observational and Prospective Study

**DOI:** 10.1101/2024.09.09.24312956

**Authors:** GA Meza-Soto, M López-González, MA Montoya-Hernández, JC Zarate-Amador, MR Galán-Solano, JA Estrada Alvarez

## Abstract

**Background:** Pediatric and adolescent cancer represents a significant public health issue nationally, being the leading cause of disease-related mortality in this age group. Previous studies in Greece, Turkey, and Mexico have shown that parents and/or caregivers have inadequate knowledge about this issue. Despite the World Health Organization’s efforts to develop new strategies, Mexico continues to experience late diagnoses. Early identification of initial signs and symptoms by parents and/or caregivers is crucial for seeking early medical attention.

**Objective:** To assess knowledge about pediatric and adolescent cancer among primary caregivers.

**Methods:** An observational, descriptive, cross-sectional, and prospective study was conducted at Family Medicine Unit No.69 in Coatzacoalcos, Veracruz, from April 2023 to January 2024. With prior informed consent, the “Parent and Caregiver Survey on Knowledge of Childhood Cancer” was administered to 374 caregivers of children and adolescents aged 0 to 19 years. Qualitative and quantitative variables were analyzed using percentages and measures of central tendency, respectively. Bivariate analysis utilized the Chi-square test through SPSS software.

**Results:** A total of 374 caregivers were evaluated, with an age range of 17 to 78 years (X̅=35.5, σ=±11.8). The majority were female (60.2%), married (48.7%), maternal caregivers (54%), had professional education (38%), and were actively employed (62.3%). Of the caregivers, 77.8% exhibited inadequate knowledge and 22.2% exhibited adequate knowledge about pediatric and adolescent cancer. Statistically significant differences (p<0.05) were observed between knowledge levels and sociodemographic variables such as maternal role, age, education, marital status, and number of children.

**Conclusion:** The alternate hypothesis that caregivers at the primary level have adequate knowledge about pediatric and adolescent cancer is rejected. It is essential to develop educational strategies to enhance knowledge about childhood cancer to improve early detection, using educational materials and audiovisual media such as the OPS video “The rhythm that gives us life” along with distributing pamphlets and questionnaires aimed at caregivers.

## Introduction

Childhood and adolescent cancer is one of the leading causes of mortality in these age groups. Previous studies have shown a significant deficit in knowledge about cancer, contributing to late diagnoses and poorer prognoses. Early detection and adequate knowledge of initial signs and symptoms by parents and caregivers are crucial for improving treatment outcomes.

## Methods

An observational, descriptive, cross-sectional, and prospective study was conducted at Family Medicine Unit No. 69 in Coatzacoalcos, Veracruz, from April 2023 to January 2024. The study population included 374 caregivers of pediatric patients aged 0 to 19 years, who were evaluated using the “Survey for parents and caregivers about knowledge of cancer in children.” Informed consent was obtained from all participants. Qualitative variables were analyzed using percentages, and quantitative variables using measures of central tendency. Bivariate analysis was performed using the Chi-square test with SPSS software.

## Results

Most caregivers were female (60.2%), married (48.7%), and maternal caregivers (54%). The predominant educational level was professional (38%), and 62.3% were employed. The evaluation of knowledge about childhood cancer showed that 77.8% of caregivers had inadequate knowledge, while only 22.2% had adequate knowledge. Sociodemographic variables such as maternal caregiver role, age, education level, marital status, and number of children were significantly associated with the level of knowledge about cancer (p<0.05).

## Discussion

The primary objective of this study was to determine the knowledge of childhood and adolescent cancer among caregivers in a primary care setting. It was found that 77.8% of caregivers had inadequate knowledge, while 22.2% had adequate knowledge. These results are similar to those obtained in previous studies in Greece (Charalabopoulos et al., 2011), Turkey (Demirbag et al., 2013), and Monterrey, Mexico (Uribe et al., 2022), all of which reported a significant deficiency in knowledge among parents and caregivers. Most participants in this study were women (60%) and mothers (54%), similar to the studies in Mexico and Turkey. Despite a high educational level (38% with professional education), adequate knowledge about childhood cancer was not guaranteed, indicating a need for specific educational strategies.

## Conclusion

Knowledge about childhood cancer among caregivers of children and adolescents in primary care is variable and generally insufficient. Despite a solid educational foundation, caregivers lack adequate knowledge about childhood cancer, highlighting the need to improve awareness and understanding of this disease. It is crucial to correct misconceptions and emphasize the importance of early diagnosis through comprehensive education. Internet and social media education is a promising approach, and awareness campaigns adapted to these platforms are recommended to maximize impact. This study underscores the need for specific educational interventions to improve the health and well-being of children and adolescents.

## Data Availability

All data produced in the present work are contained in the manuscript

## Conflict of Interest

The authors declare no conflicts of interest related to this study.

## Acknowledgements

We thank all the caregivers who participated in the study and the staff of Family Medicine Unit No. 69 for their collaboration.

---

**Figure 1.**
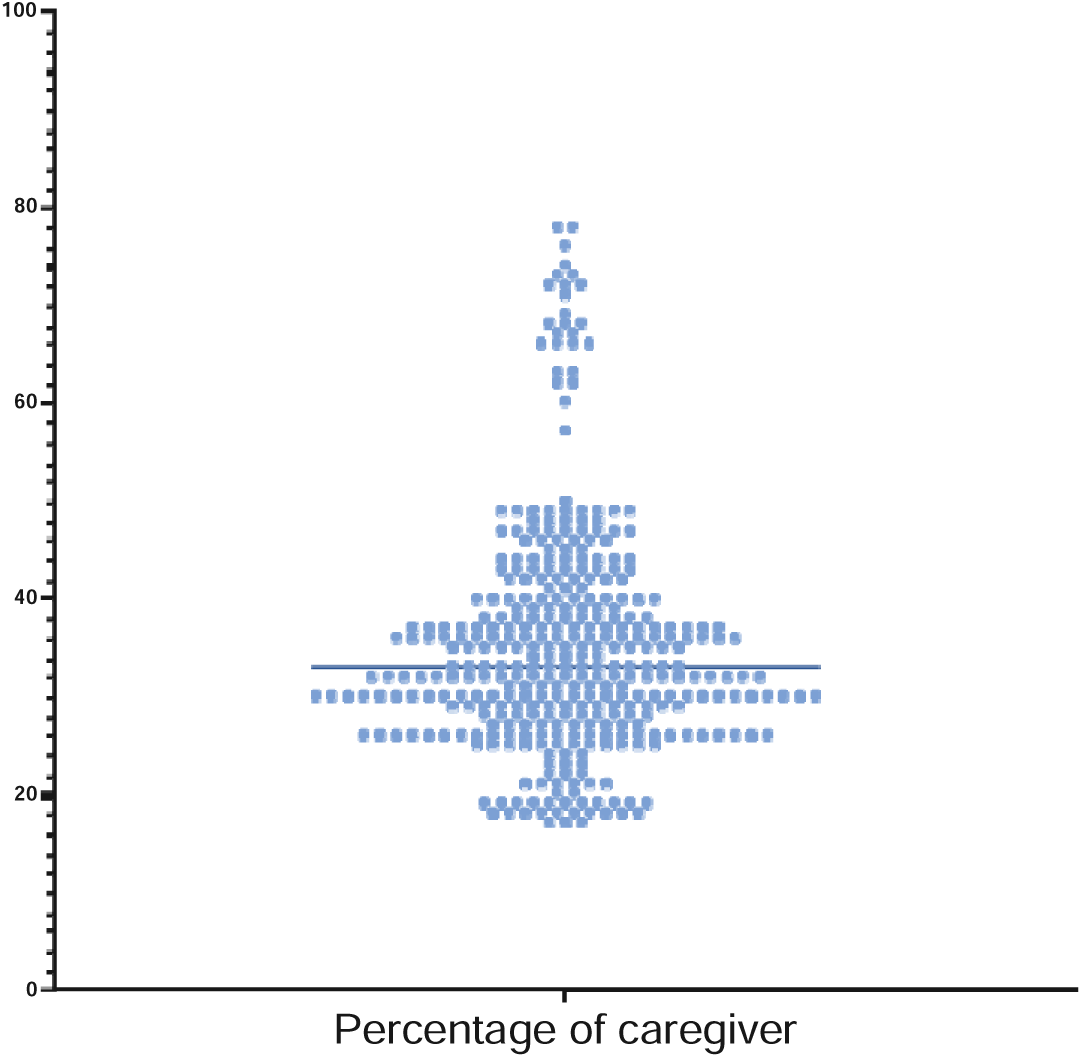
Age distribution of patients. Age distribution with XLJ of 35.35 and σ of 11.81 years.

**Figure 2.**
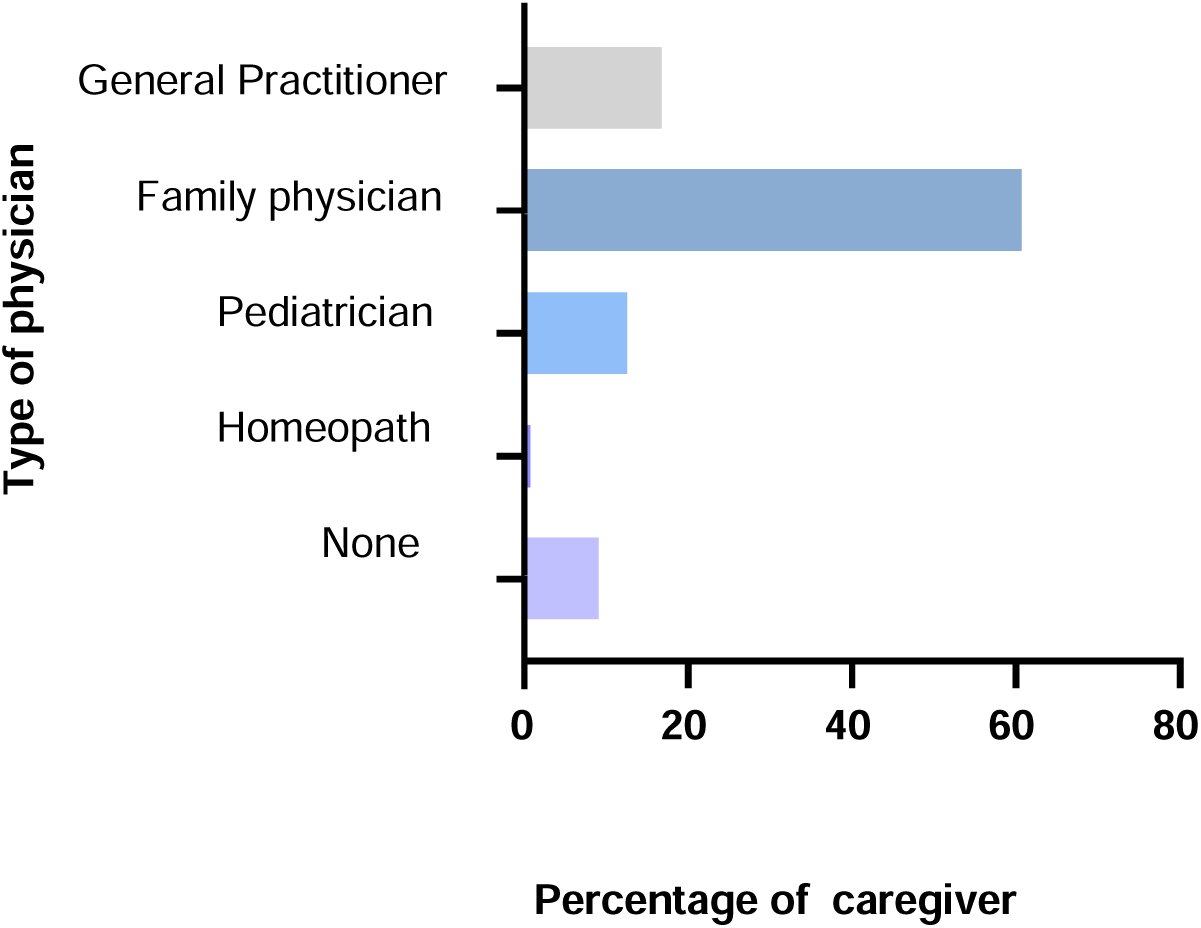
Consulting physician. The physician with the highest annual visits who is responsible for assessing children and adolescents is the family physician.

**Figure 3.**
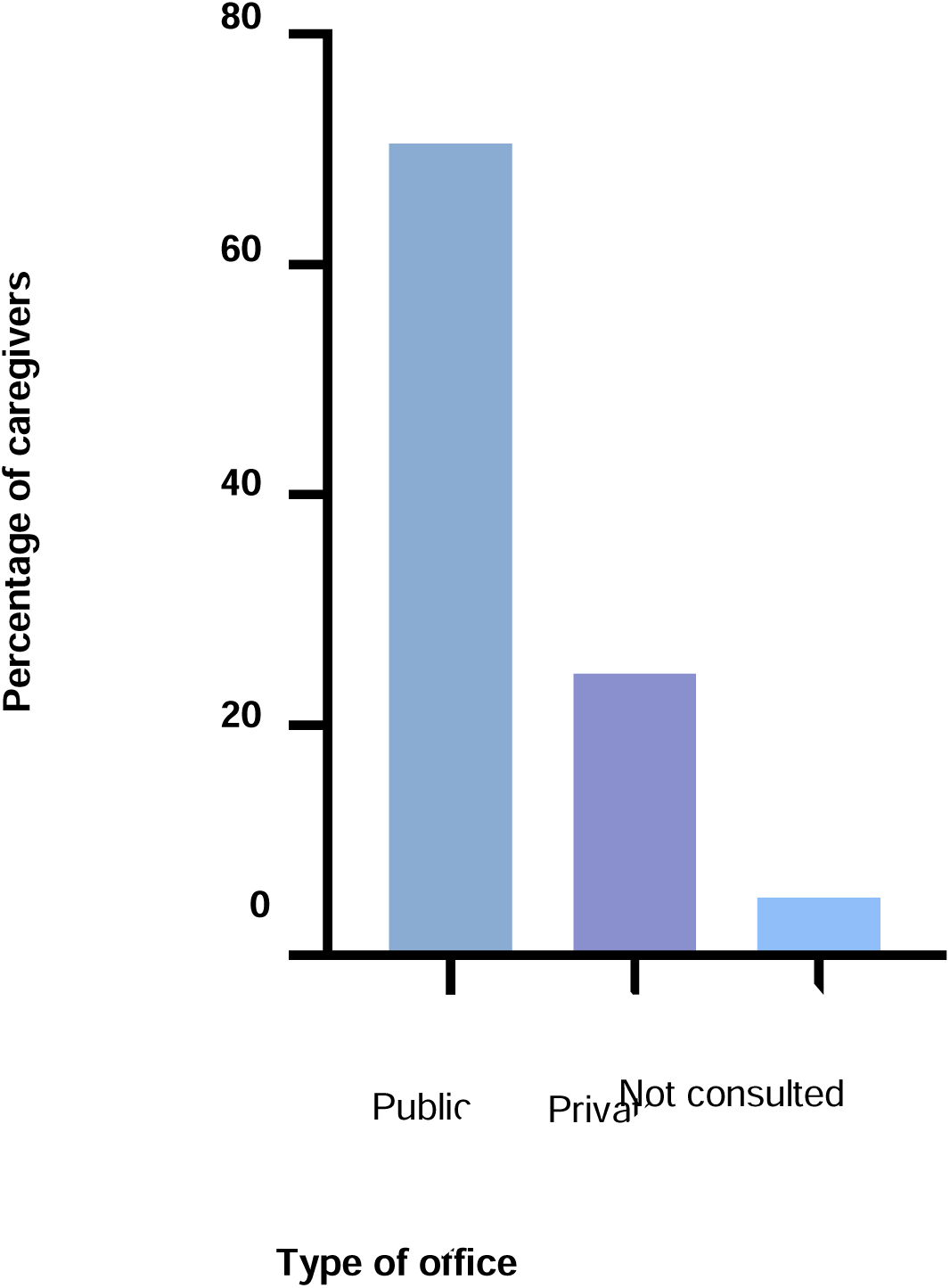
Visited office. Caregivers report taking children and adolescents for evaluation to the public office in 70.5% of cases

**Figure 4.**
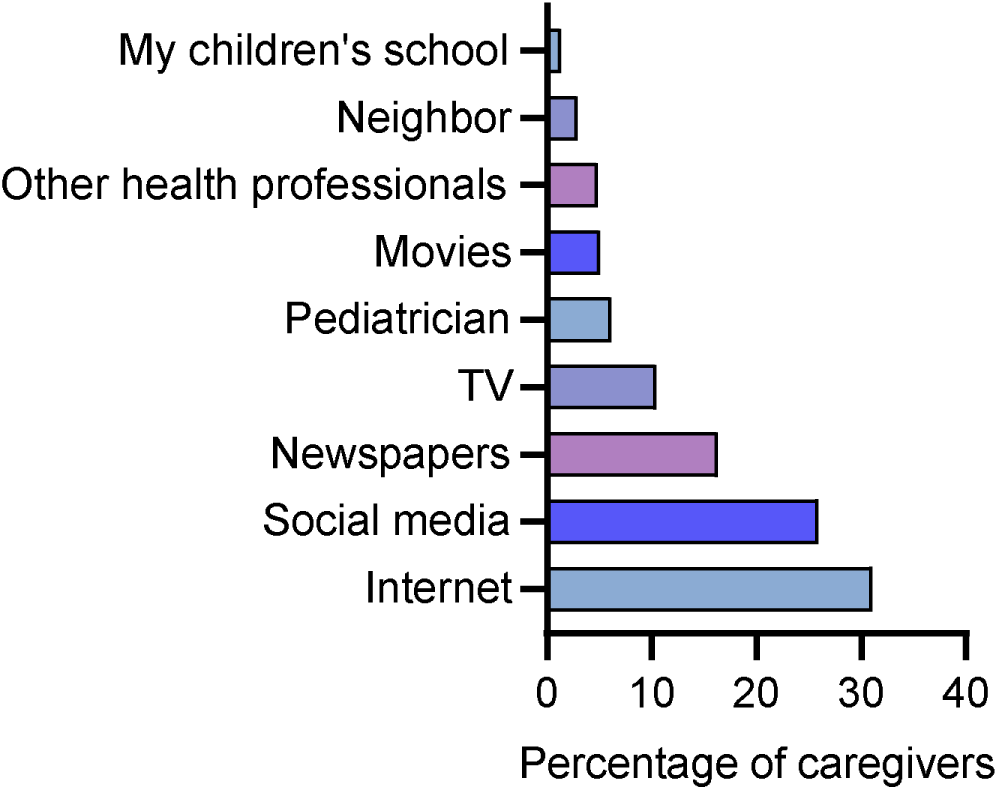
Source of information on childhood cancer. The sources of information from which caregivers have gained the most knowledge are the internet, social media, and newspapers.

**Figure 5.**
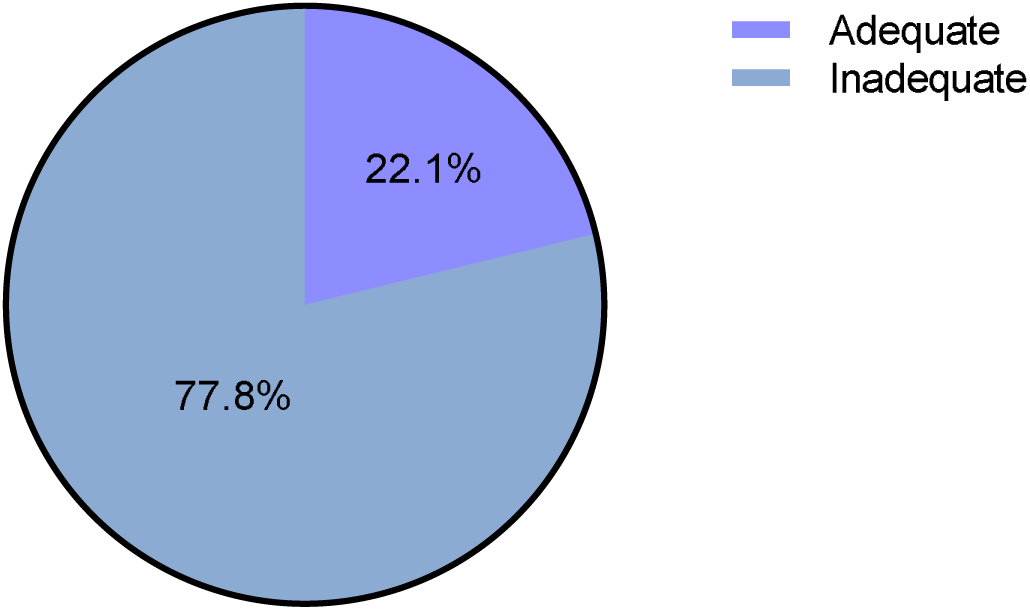
Knowledge about childhood cancer. 77.8% exhibited inadequate knowledge about childhood cancer.

**Table 1.**
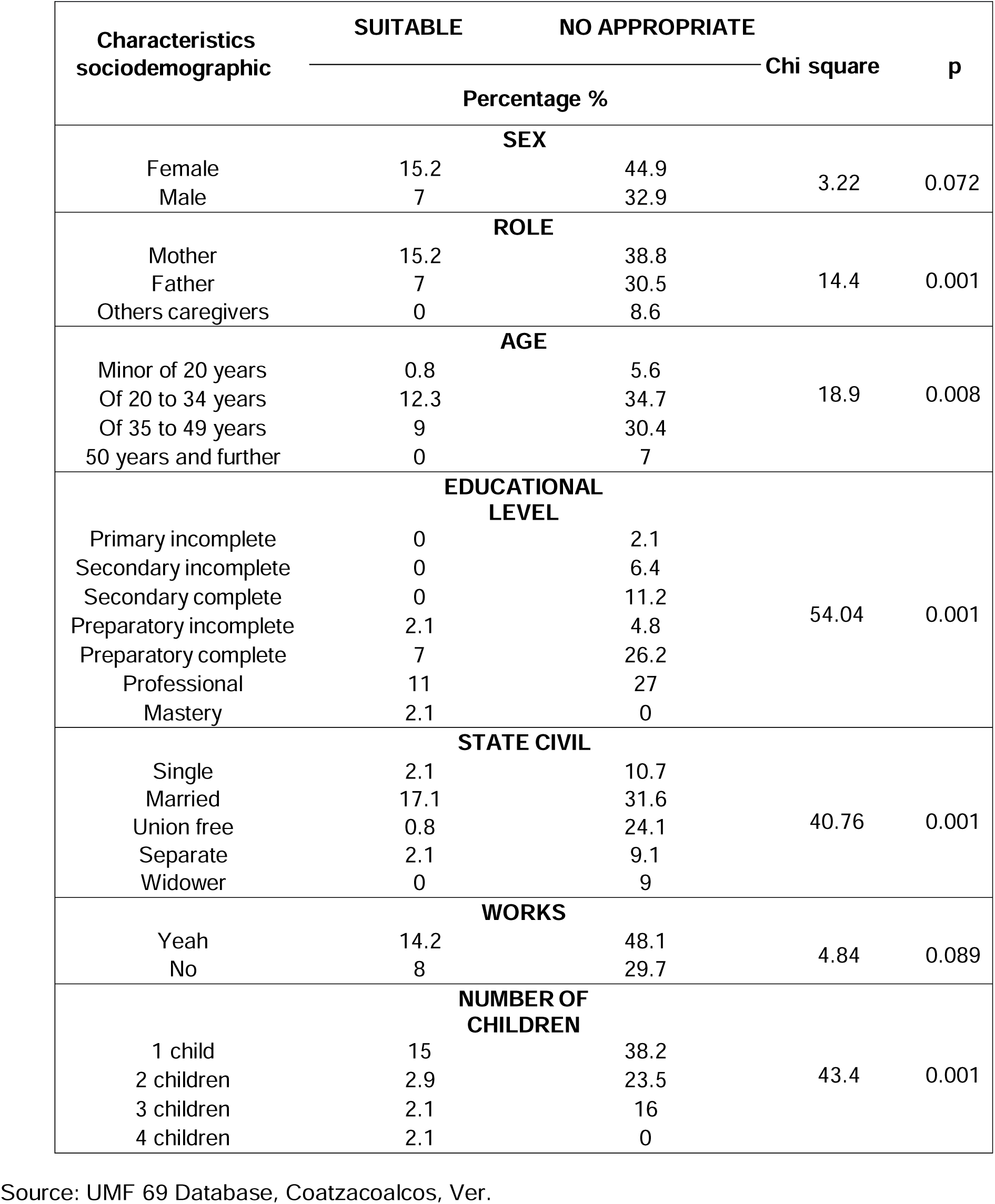
Analysis of knowledge about cancer childhood and adolescent cancer in caregivers associated with demographic characteristics.

**Table 2.**
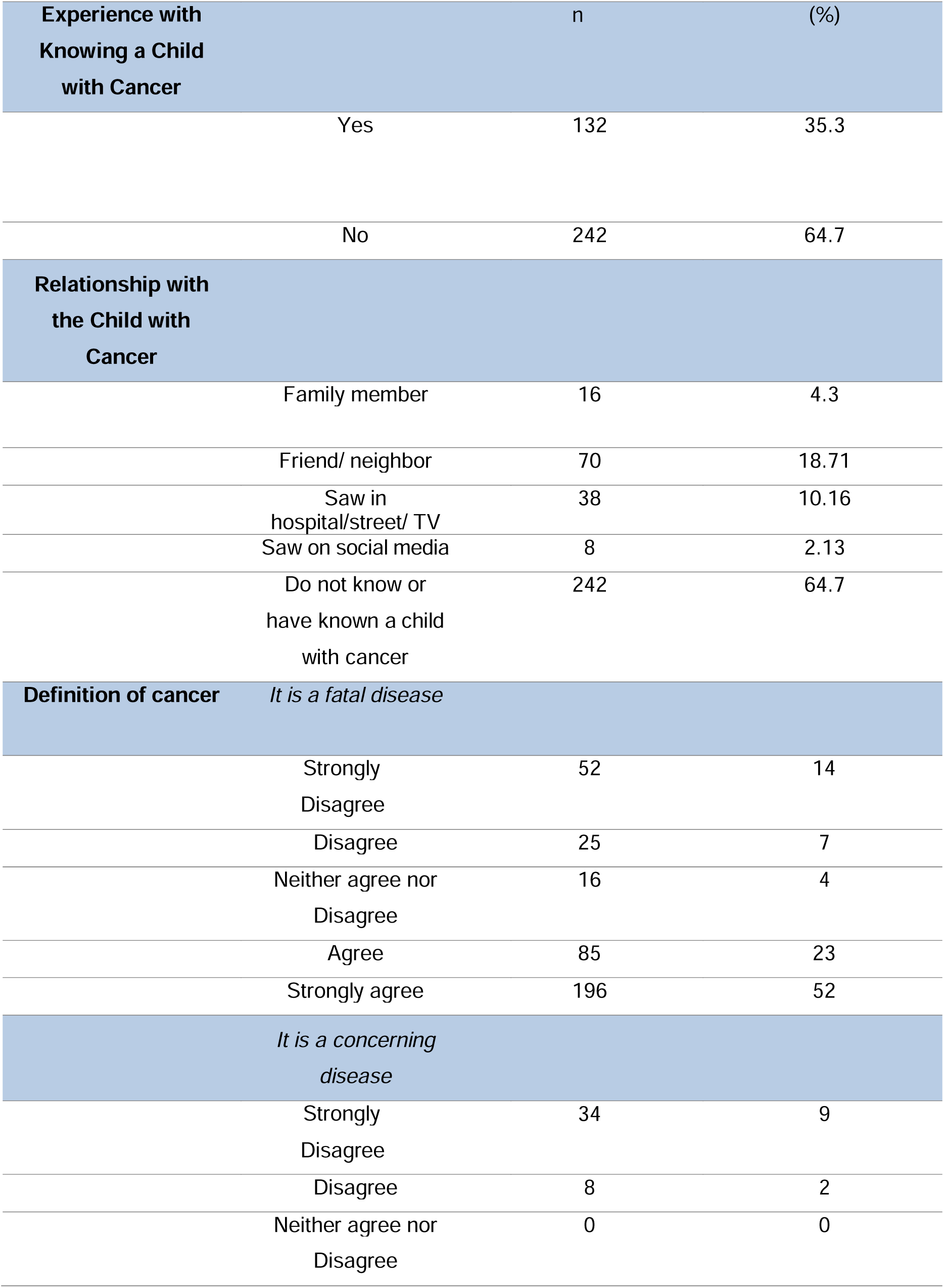
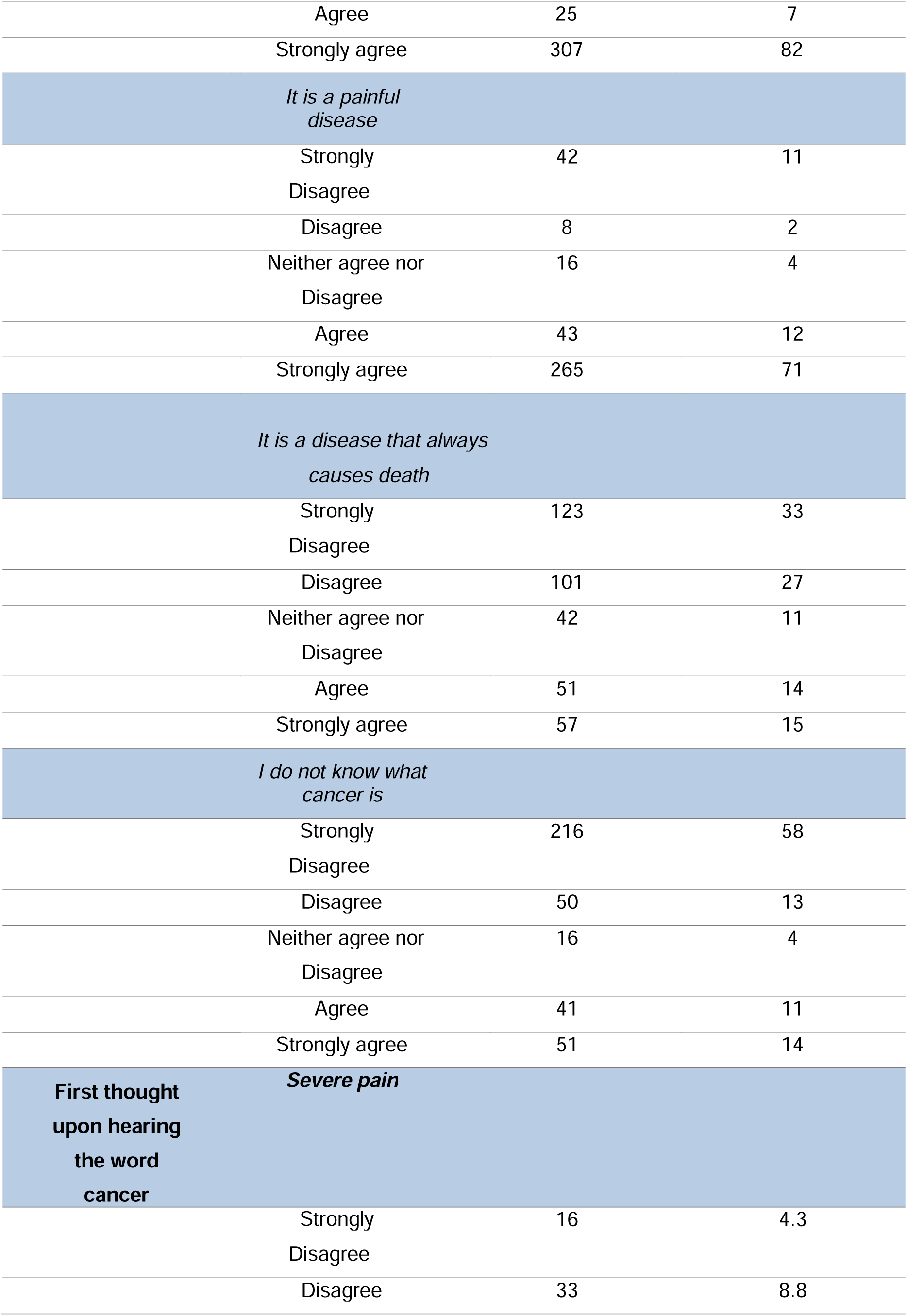
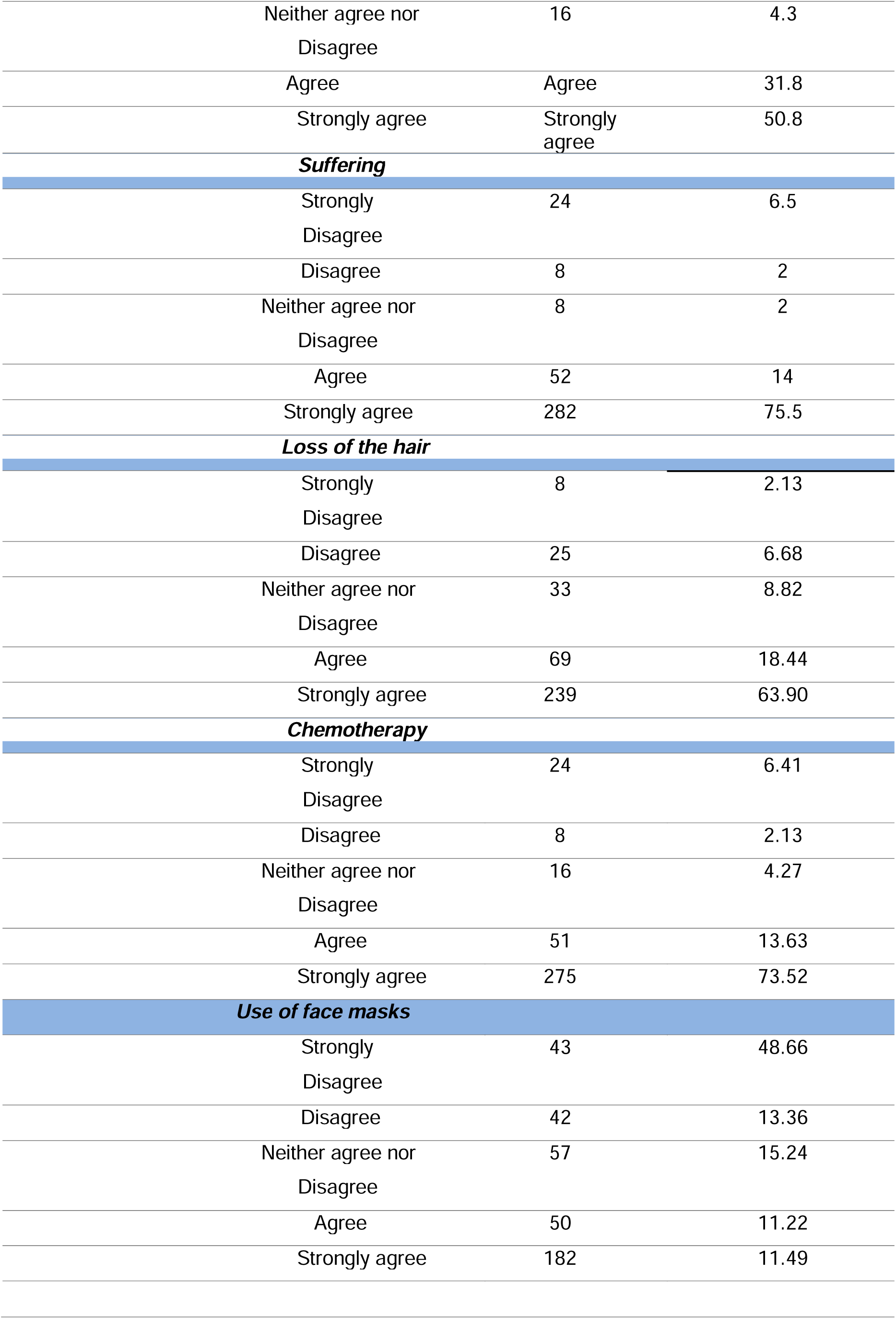
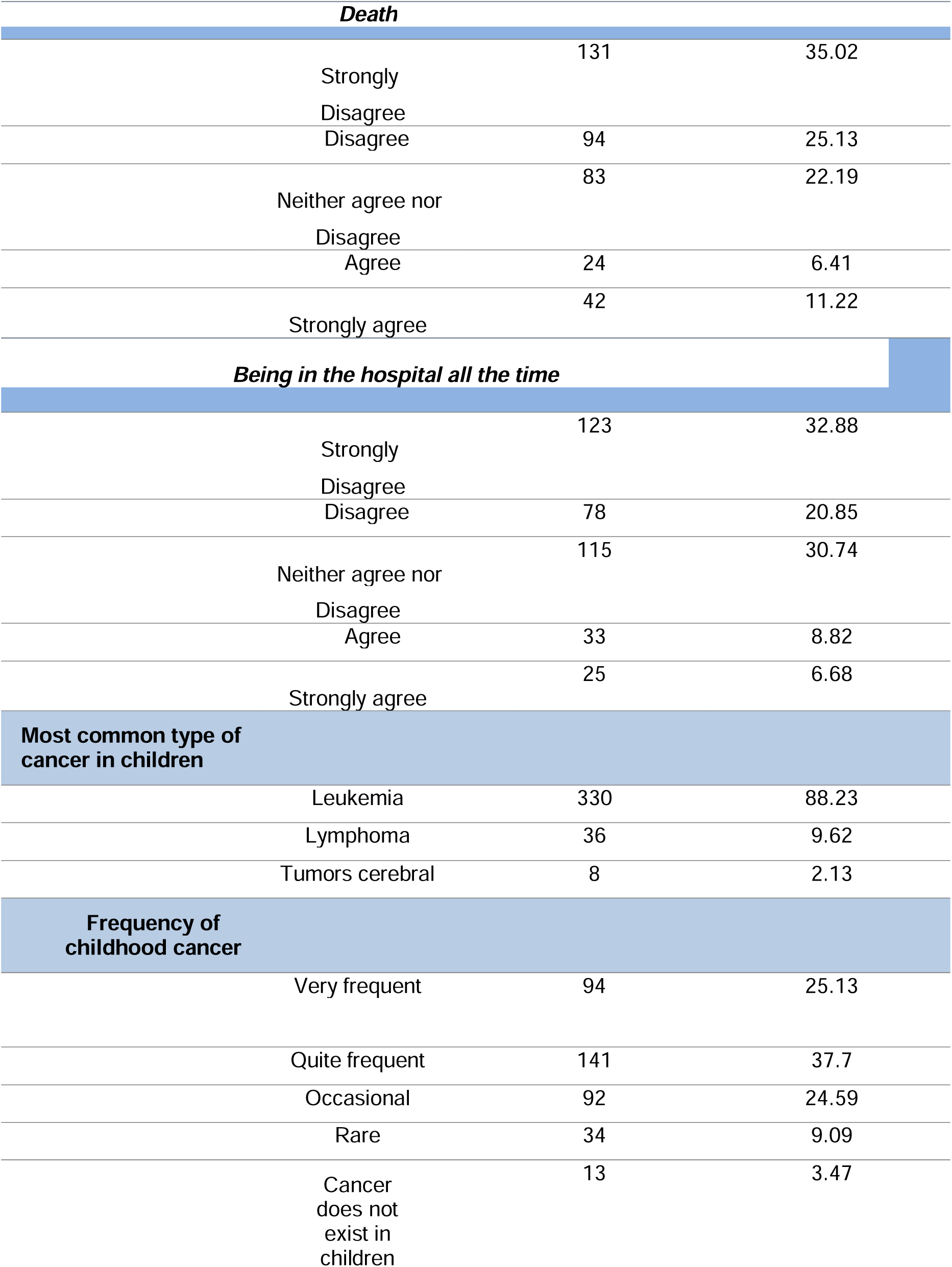
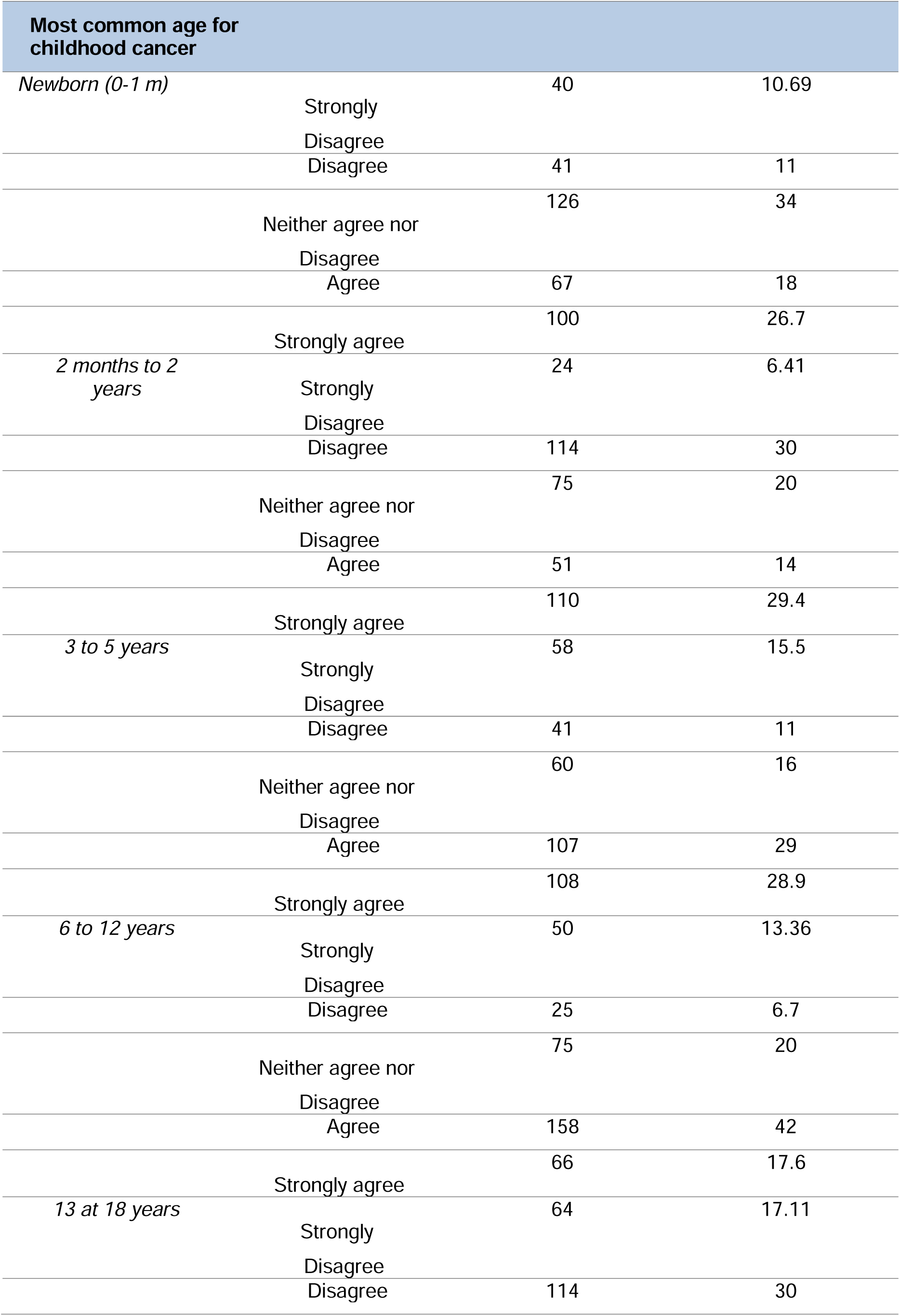
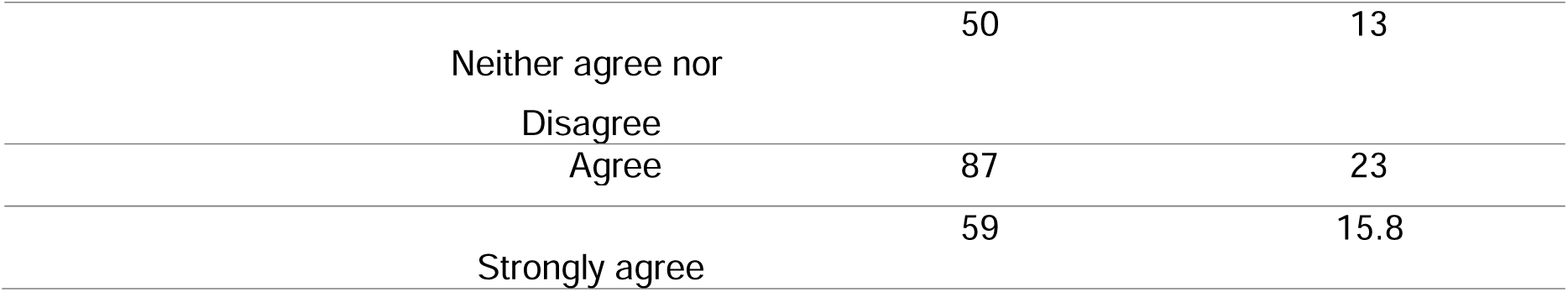
Participant Background on Knowledge of a Child with Cancer, Relationship with the Child, and Knowledge about the Cancer. This table first associates the caregivers’ relationship with a child with cancer and the knowledge they reported about childhood and adolescent cancer, related to its definition and epidemiology

**Table 3.**
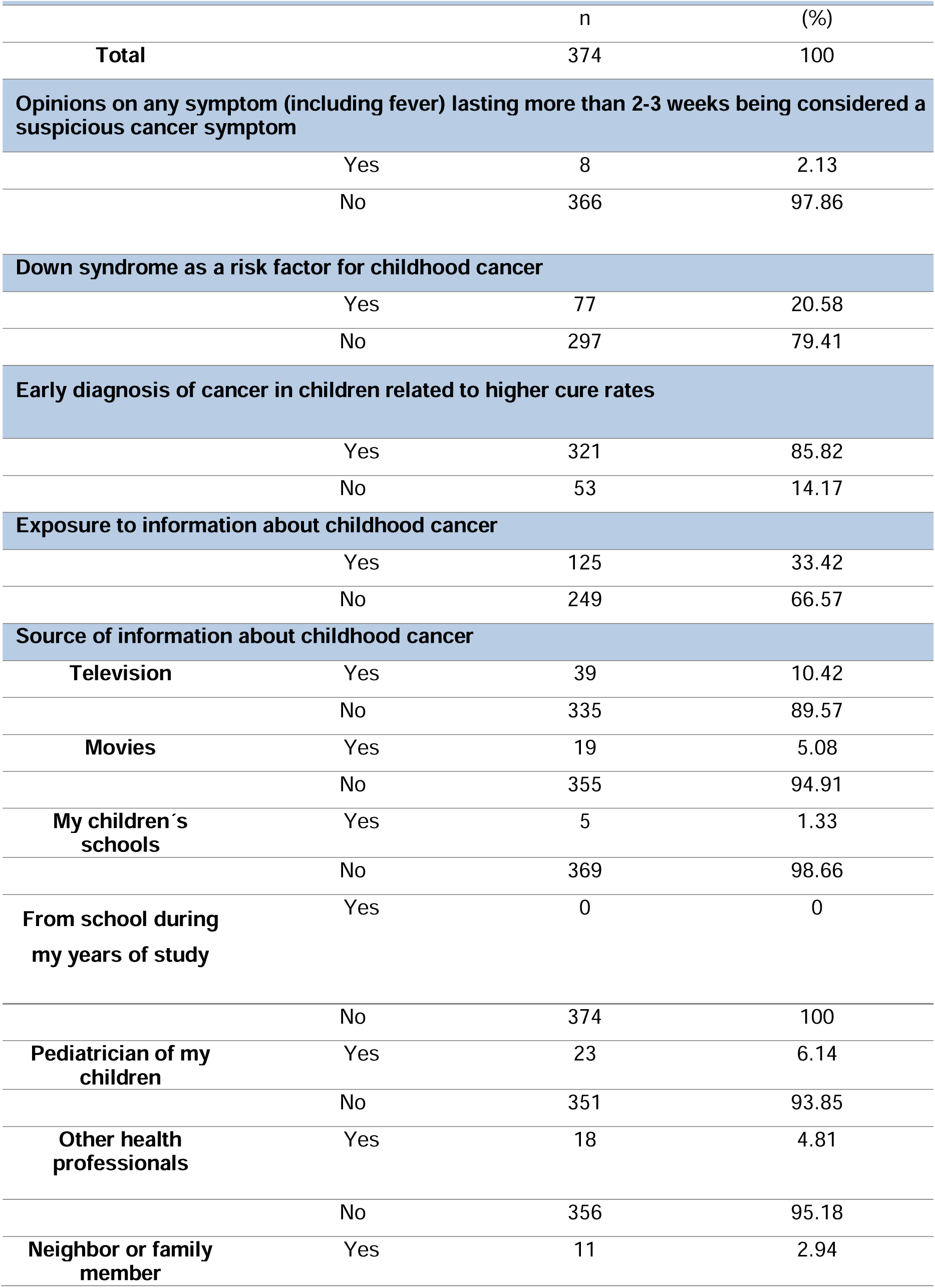
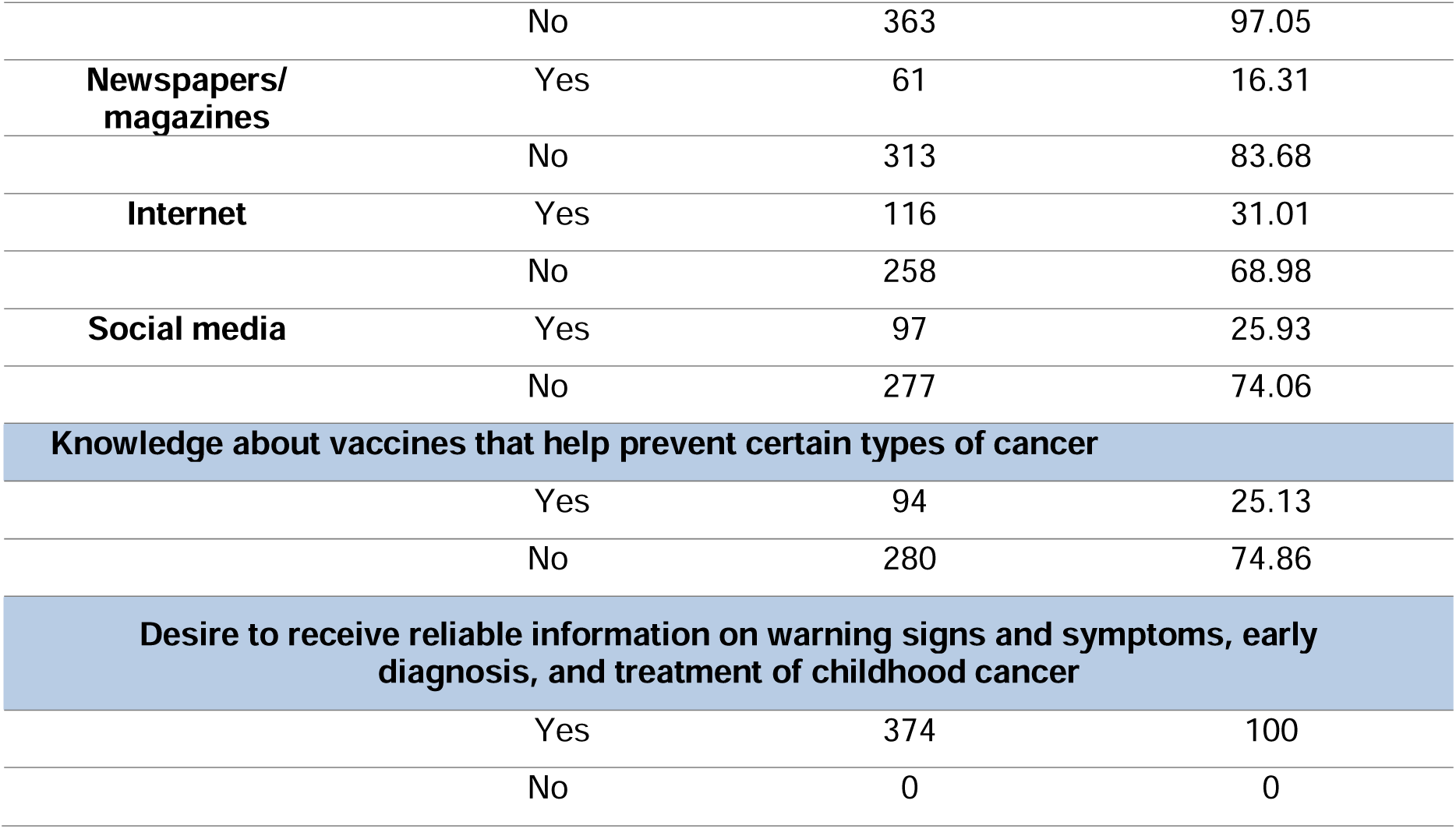
Parents’ and Caregivers’ Statements on Symptoms, Early Diagnosis Strategies, and Exposure to Information about Childhood Cancer. This table shows caregivers’ perceptions regarding symptoms, risk factors, and sources of information about childhood cancer.

## Notes

### Competing Interest Statement

The authors have declared no competing interest.

### Funding Statement

This study did not receive any funding

### Author Declarations

The ethics committee of the Mexican Institute of Social Security / General Hospital of Zone No. 32 approved the ethical aspects of this work.

## Bibliographic references

1. Tinoco A. Definición de cáncer: una controversia científica entre el paradigma ortodoxo y el crítico en oncología. Rev. colomb filos cienc [Internet]. 4 de Octubre de 2019;19(38). Available at: 10.18270/rcfc.v18i36.2271

2. Rivera R. La importancia del cáncer infantil en México. Gac Mex Oncol [Revista en la Internet].2022;21(1-2). Available at: 10.24875/j.gamo.m22000218

3. Arenas Á, Torrado E, Garrido M. Intervención familiar en diagnóstico reciente e inicio de tratamiento del cáncer infantil. Apunt Psicol [Internet]. 2022;213–20. Available at: 10.55414/ap.v34i2-3.612

4. Who.int. El cáncer infantil. [Consultado el 1 de abril de 2022]. Available at: https://www.who.int/es/news-room/fact-sheets/detail/cancer-in-children.

5. Childhood cancer of the Brain and Other Nervous System - Cancer stat facts [Internet]. SEER. [Consultado el 3 de abril de 2022]. Available at: https://seer.cancer.gov/statfacts/html/childbrain.html

6. National Cancer Institute. Cancer of Any Site-Cancer Stat Facts [Internet]. SEER. 2018.[Consultado 3 de abril 2022] Available at: https://seer.cancer.gov/statfacts/html/all.html

7. Handayani K, Sitaresmi M, Supriyadi E, Widjajanto P, Susilawati D, Njuguna F, et al. Delays indiagnosis and treatment of childhood cancer in Indonesia. Pediatric Blood & Cancer. 2016 Aug 11;63(12):2189–96. Available at: 10.1002/pbc.26174

8. Arora R, Kanwar V, Manglani M, Bhat S, Banavali S, Raj R, et al. WHO Global Initiative for childhood cancer-India responds. Pediatric Hematology Oncology Journal. [Internet] 2020 Sep. [Consultado 5 abril de 2022] Vol.5 (4):145–150. Available at: 10.1016/j.phoj.2020.06.005

9. Bazán U, Pontificia Universidad Católica del Perú. Análisis de la Nueva Ley de Urgencia Médica para la Detección Oportuna y Atención Integral del Cáncer del Niño y del Adolescente. peryfa [Internet]. 2020;(9):17–33. Available at: 10.33539/peryfa.2020.n9.2332

10. Gil M, Pacheco D, Santos F, Villasís M, Betanzos Y, Miranda G. Early discharge of pediatric patients with cancer, fever, and neutropenia with low-risk of systemic infection. Bol Med Hosp Infant Mex [Internet]. 2018;75(6):352–7. Available at: 10.24875/BMHIM.18000015

11. GBD 2017 Childhood Cancer Collaborators. The global burden of childhood and adolescent cancer in 2017: an analysis of the Global Burden of Disease Study 2017. Lancet Oncol [Internet]. 2019;20(9):1211–25. Available at: 10.1016/S1470-2045(19)30339-0

12. Dorantes E, Ávila D, Klünder M, Juárez L, Márquez H. Survival in pediatric patients with cancer during the COVID-19 pandemic: scoping systematic review. Bol Med Hosp Infant Mex [Internet]. 2020;77(5):234–41. Available at: 10.24875/BMHIM.20000174

13. Arsenault V, Qiu W, Liu Q, Yeh J, Leisenring W, Ness K, et al. Emergency department (ED) visits and hospitalizations in survivors of childhood cancer in the Childhood Cancer Survivor Study. J Clin Oncol [Internet]. 2019;37(15_suppl):10056–10056. Available at: 10.1200/jco.2019.37.15_suppl.10056

14. Vásquez L, Silva J, Chávez S, Zapata A, Diaz R, Tarrillo F, et al. Prognostic impact of diagnostic and treatment delays in children with osteosarcoma. Pediatr Blood Cancer [Internet]. 2020;67(4):e28180. Available at: 10.1002/pbc.28180

15. Russo A, Blettner M, Merzenich H, Wollschlaeger D, Erdmann F, Gianicolo E. Incidence of childhood leukemia before and after shut down of nuclear power plants in Germany in 2011: A population-based register study during 2004 to 2019. Int J Cancer [Internet]. 2022; [Consultado 7 abril de 2022; Vol.152(5):913–920. Available at: 10.1002/ijc.34303

16. Uribe L, Garza B, Vázquez A, Castorena F, Rodríguez J. Exploring knowledge of parents and caregivers on cancer symptoms in children: an observational study regarding the need for educational tools and health promotion in low- and middle-income countries. BMC Pediatr (Internet). 2022;22(1):638. Available at: 10.1186/s12887-022-03686-4.

17. Liu J, Shanmugavadivel D, Callcut A, Taylor E, Walker D. 1195 Childhood, teenagers and young adult cancer symptom awareness amongst future doctors. At: Abstracts. BMJ PublishingGroup Ltd and Royal College of Paediatrics and Child Health; 2021.

18. Weyl Arush M, Haimi M, Peretz M, Rennert H, Katz K. 640 Delay in diagnosis of children withcancer: a retrospective study of 315 children. EJC Suppl [Internet]. 2003;1(5):S192. Available at: 10.1016/s1359-6349(03)90672-0

19. Schraw J, Desrosiers T, Nembhard W, Langlois P, Meyer R, Canfield M, et al. Cancer diagnostic profile in children with structural birth defects: An assessment in 15,000 childhood cancer cases. Cancer [Internet]. 2020;126(15):3483–92. Available at: 10.1002/cncr.32982

20. Dixon S, Chen Y, Yasui Y, Pui C, Hunger S, Silverman L, et al. Impact of risk-stratified therapy on health status in survivors of childhood acute lymphoblastic leukemia: A report from the childhood cancer survivor study. Cancer Epidemiol Biomarkers Prev [Internet]. 2022;31(1):150– 60. Available at: 10.1158/1055-9965.EPI-21-0667

21. Karimi M, Mehrabani D, Yarmohammadi H, Jahromi FS. The prevalence of signs and symptoms of childhood leukemia and lymphoma in Fars Province, Southern Iran. Cancer Detect Prev [Internet]. 2008;32(2):178–83. Available at: 10.1016/j.cdp.2008.06.0016

22. Zinser J. Tabaquismo y cáncer de pulmón. Salud Publica Mex [Internet]. 2019;61(3):303–7. Available at: 10.21149/10088

23. González M, Bermeo J, Laverde L, Tafurt Y. Carcinógenos ambientales asociados a cáncer infantil. Univ salud [Internet]. 019;21(3):270–6 Available at: 10.22267/rus.192103.164

24. Steliarova E, Colombet M, Ries L, Moreno F, Dolya A, Bray F, et al. International incidence of childhood cancer, 2001–10: a population-based registry study. Lancet Oncol [Internet]. 2017;18(6):719–31. Available at: 10.1016/s1470-2045(17)30186-9

25. Andrade BP, Wolff C, Saldanha FL, Oliveira GMSF de, Lima IB de AL, Nelson LA de S, et al. Diagnóstico precoce no câncer infantil como estratégia para garantir qualidade de vida. At: Medicina: Ciências da saúde e pesquisa interdisciplinar 3. Atena Editora; 2021. p. 65–72.

26. Ferrari A, Lo Vullo S, Giardiello D, Veneroni L, Magni C, Clerici C, et al. The sooner the better? How symptom interval correlates with outcome in children and adolescents with solid tumors: Regression tree analysis of the findings of a prospective study: Delay in diagnosis and outcome. Pediatr Blood Cancer [Internet]. 2016;63(3):479–85. Available at: 10.1002/pbc.25833

27. Zapata M, González E, Doubova S, Menendez N, Cruz C, Gonzalez R, et al. Patient and health service factors associated with delays in cancer treatment for children without social security in Mexico. Pediatr Blood Cancer [Internet]. 2020;67(9):e28331. Available at: 10.1002/pbc.28331

28. Friestino J, Corrêa C, Filho D de C. Percepções dos Profissionais sobre o Diagnóstico Precoce do Câncer Infantojuvenil na Atenção Primária à Saúde. Rev BrasileiraDeCancerologia [Internet]. 2019;63(4):265–72. Available at: 10.32635/2176-9745.rbc.2017v63n4.127

29. Dragomir M. OC-85 Early diagnosis in chilhood cancer saves lives. At: Oral Communications. BMJ Publishing Group Ltd and Royal College of Paediatrics and Child Health; 2017.

30. Wang X, Brown D, Cao Y, Ekenga C, Guo S, Johnson K. The impact of health insurance coverage on racial/ethnic disparities in US childhood and adolescent cancer stage at diagnosis. Cancer [Internet]. 2022;128(17):3196–203. Available at: 10.1002/cncr.34368

31. Godono A, Felicetti F, Conti A, Clari M, Dionisi M, Gatti F, et al. Employment among childhoodcancer survivors: A systematic review and meta-analysis. Cancers (Basel) [Internet]. 2022;14(19):4586. Available at: 10.3390/cancers14194586

32. Mulrooney D. Challenges predicting the cardiovascular future for survivors of childhood cancer. Cancer Epidemiol Biomarkers Prev [Internet]. 2022;31(3):515–6. Available at:: 10.1158/1055-9965.EPI-21-1329

33. 15 de febrero, Día Internacional del Cáncer Infantil [Internet]. Insp.mx. [citado el 9 de diciembre de 2022]. Available at: https://www.insp.mx/avisos/15-de-febrero-dia-internacional-del-cancer-infantil

34. Zayas A, Sarahí J. Conocimiento y concientización sobre el cáncer Infantil en estudiantes demedicina de México. 2020 [citado el 9 de diciembre de 2022];643574. Available at: https://repositorio.tec.mx/handle/11285/643574

35. Caballero N. Cultura e inteligencia musical mediante la investigación como estrategia pedagógica en educación básica. Cult Educ Soc [Internet]. 2018;9(3):813–22. Available at: 10.17981/cultedusoc.9.3.2018.96

36. Public Health Agency of Canada, Statistics Canada, Canadian Cancer Society, provincial/territorial cancer registries. Release notice - Canadian cancer statistics 2019. Health Promot Chronic Dis Prev Can [Internet]. 2019;39(8–9):255. Available at: 10.24095/hpcdp.39.8/9.04

37. Gharehzadehshirazi A, Zarejousheghani M, Falahi S, Joseph Y, Rahimi P. Biomarkers and corresponding biosensors for childhood cancer diagnostics. Sensors (Basel) [Internet]. 2023;23(3):1482. Available at: 10.3390/s23031482

38. Dewan P, Jain P, Trivedi M. Childhood Cancer in India: Miles to Go Before We Sleep!. [On line] Indian Pediatr. 2023 Dec 15;60(12):1032–1034. https://www.indianpediatrics.net/dec2023/1032.pdf

39. Charalabopoulos K, Makris G, Charalabopoulos A, Golias C, Athanasiou K. Public knowledge, beliefs and practices in Greece about cancer etiology and prevention. East Mediterr Health J [Internet]. 2011;17(05):392–7. Available at: 10.26719/2011.17.5.392

40. Demirbag BC, Kurtuncu M, Guven H. Knowledge of Turkish mothers with children in the 0-13 age group about cancer symptoms. Asian Pac J Cancer Prev [Internet]. 2013;14(2):1031–5. Available at: 10.7314/apjcp.2013.14.2.1031

